# Resolutive results with oral corticosteroids for patients with COVID-19 in pulmonary inflammatory phase. Successful outpatient experience during the collapse of Belém do Pará Health System - Brazil

**DOI:** 10.1101/2021.04.19.20219949

**Authors:** Roberto S. Zeballos, Luciana N.L. Cruz, Vânia C.R. Brilhante, V. Lima Adriana, Carlliane L.L.P. Martins, Karla N.A. Lima, Michele C.G.L. Leão, Belinete L. Cruz, Alessandra B. Dysarz, Paulo C.B. Lobato, Erika S. Almeida, Rute A.P. Costa, Klevison C. Araújo, Augusto C.C. Almeida, Leônidas B.D. Junior, Kathia O. Harada, Rita H.V.S. Araújo, Elaine X. Prestes, Ivete M.S. Souza, Avany M.P. Pereira, Antônio B.M.A. Rocha, Iracema A. Estevão, Fagnei I.C. Carvalho, Paulo M.P. Melo

## Abstract

In the face of new diseases, medicine needs to reinvent itself in order to contain and control epidemics, such as the one we have recently faced, COVID-19, a disease with a wide spectrum of clinical severity. <u>A new moment has been established, since the application of well known, effective and safe medications for other diseases, has shown high success rates in the treatment of COVID-19.</u> Thereunto, studies with early intervention are needed, which can change the unfavorable outcome of patients. In this article, we report the successful experience using an oral strategy during the collapse of Belém do Pará Health System, Brazil. Two hundred and ten patients were diagnosed with respiratory failure due to COVID-19, with no option of hospital treatment due to lack of beds and resources. These patients were then started on therapeutic regimen consisting of prednisolone, enoxaparin and macrolides associated and followed in outpatient facilities. Two hundred and eight patients had excellent therapeutic response and there were only two fatalities. These results push research boundaries, valuing outpatient treatment with early use of prednisolone in the initial pulmonary phase, preventing severe COVID-19 pneumonitis. Adoption of the proposed treatment intends to reduce the need for hospitalization, as well as lethality, with social robust benefits and incalculable economic savings since involves the use of accessible, safe and not expensive medications.

## INTRODUCTION

COVID-19 is a disease with a wide spectrum of clinical severity, from asymptomatic to fatal. In the absence of clinical trials and guidelines, with hospitalizations and increasing mortality, often in a chaotic scenario that compromises the effectiveness of actions, it is necessary to create therapeutic strategies based on pathophysiological knowledge (1). Furthermore, the speed of spread of COVID-19 is surprising, generating numerous simultaneous infections. 10 to 15% of patients evolve with greater severity, requiring medical care and supplemental oxygen, and another 5% evolve into a critical condition with the need for intensive care due to respiratory failure, septic shock, thromboembolism and multiple organ dysfunction (2).

In Europe, there were already reports of successful experiences in reducing hospital mortality with the use of corticosteroid pulse therapy, confirming that treatment with methylprednisolone may be beneficial for patients who develop acute respiratory distress syndrome (ARDS) in the disease progression (3). Corticosteroids were discovered in 1935 and approved by the Food and Drug Administration in the 1950s. Supplied in many different formulations, they are widely used on treatment of a variety of diseases for its anti-inflammatory and immunosuppressive effects (Kapugi, 2019). In Brazil, inspired by an article published in JAMA (4) by Chaomin et al, we adopted a inpatient, lower, antiinflammatory doses of intravenous methylprednisolone, in São Paulo, with excellent results since March 2020. However, hospital efforts in an attempt to introduce corticosteroid treatment in the inflammatory phase, even before randomized controlled studies confirm its benefit (5), would not be able to contain the worsening of the disease and deaths due to the high amount of simultaneous patients seeking hospital care, since there were lack of hospital structure in the scenario of collapse.

In order to reduce the pressure on hospital care and also to reduce morbidity and mortality, we introduced a resolutive solution with an oral therapeutic scheme in the face of emergency circumstances in the collapse of the health system in the city of Belém do Pará.

## CASUISTRY AND METHODS

From April 31 to May 15, 210 patients in inflammatory pulmonary phase were treated with the protocol adapted to the situation of the collapse of the health system in the metropolitan region of Belém, capital of the state of Pará, Brazil, whose characteristics are exposed in Figure 1 (Figure 1). It is essential to understand that the collapse of the public and private health system made it impossible for the study patients to receive hospital medical care, despite the clinical indication. During the intervention period, the emergency room’s capacity was exhausted and hospitals were literally closed due to lack of free hospital beds.

**ANEXOS – FIGURE 1.**
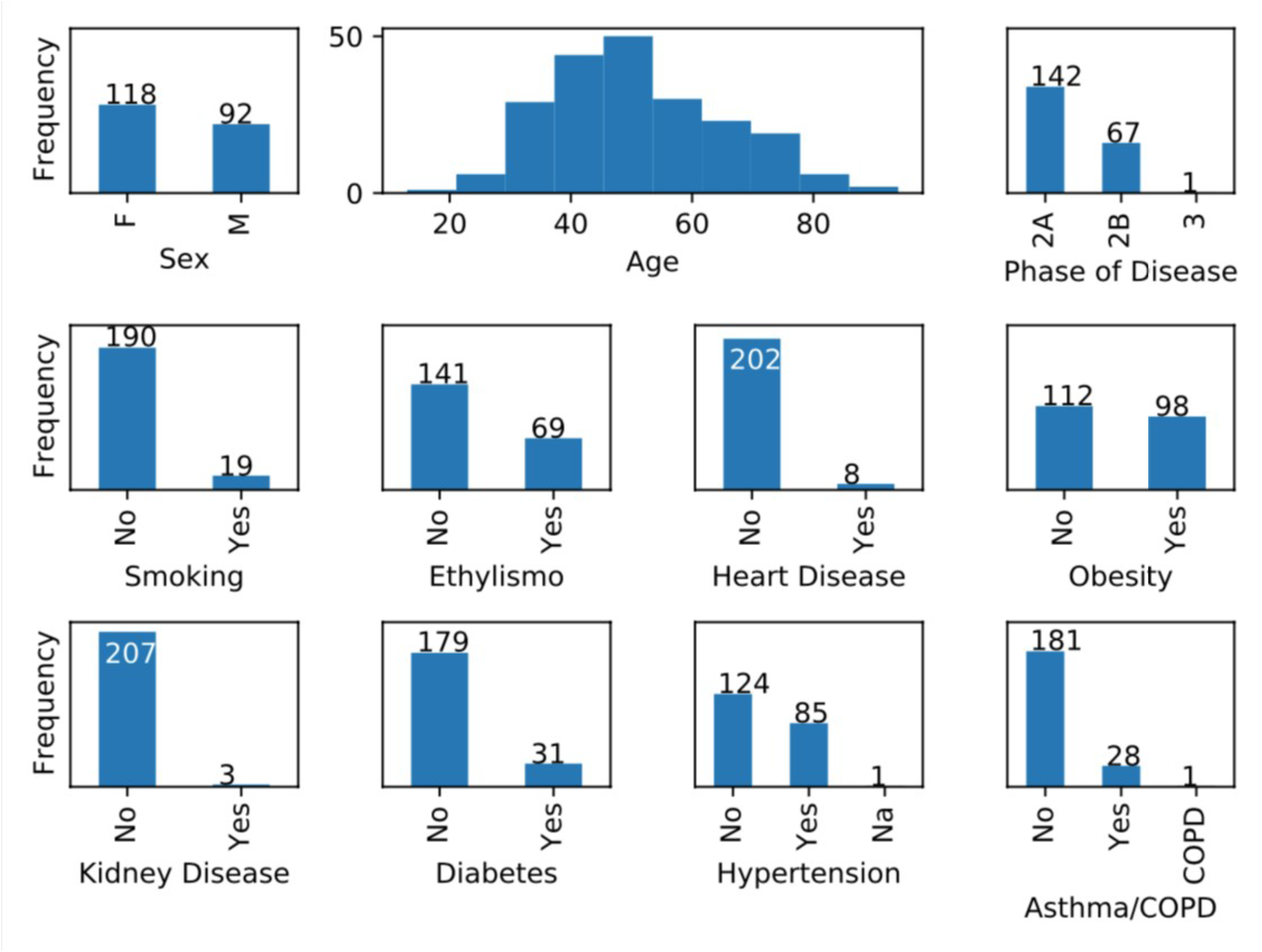
CHARACTERISTICS OF THE STUDY POPULATION.

The criteria used for inclusion in the study and the methods of clinical and diagnostic evaluation, as well as the therapeutic scheme used are listed below.

### INCLUSION CRITERIA FOR ORAL THERAPEUTIC STRATEGY

1. Affected by COVID-19;
2. Stage II pulmonary inflammation, according to SIDDIQI classification (6) with respiratory failure;
3. Absence of pulmonary fungal infection;
4. Hospital structure unavailable for treatment.

### CLINICAL AND DIAGNOSTIC EVALUATION METHODS

1. RT-PCR (reverse transcription followed by polymerase chain reaction) when available;
2. Specific serology;
3. D-dimer when available;
4. Chest Computed Tomography - Classification of CO-RADS (7) and divided into quartiles (<25%, between 25 and 50%, between 50 and 75%,> 75%);
5. Clinical condition of respiratory failure with at least one of the criteria above changed.

### ORAL THERAPEUTIC STRATEGY

The medications used were adapted from the study design of Dr. Roberto Zeballos with Dr. Marcelo Amato for patients hospitalized with methylprednisolone, enoxaparin, clarithromycin and Ceftriaxone.

Thus, we carry out the following therapeutic scheme (adaptation for oral treatment):

1. Prednisolone 40 mg once a day for 7 days;
2. Clarithromycin 500 mg once a day for 7 days;
3. Enoxaparin 40 mg SC once a day for 7 days;
4. Axetylcefuroxime 500 mg every 12 hours for 7 days.

We used alternative antibiotics such as levofloxacin or moxifloxacin, considering the risk of shortages in pharmacies and also the social and economic situation of the study patients. Supportive measures such as pronation, supplemental oxygen and respiratory physiotherapy were used when available and necessary.

## RESULTS AND DISCUSSION

Of the 210 cases, only two deaths occurred. As far as we know, this study represents the first large series of cases of patients with severity criteria who were treated at home, considering the pathophysiological rationale, given the collapse of the health system in the metropolitan region of Belém-Pará-Brazil. The outpatient treatment proposed in the study showed that it was decisive in the clinical improvement of patients who were progressing severely or were already in a severe state, with only two deaths. Even among patients who maintained a serious condition, home treatment stabilized the patient and ensured his survival until hospitalization could be feasible. Once indicated with criteria and monitoring, this home treatment may be able to reduce pressure for hospital care, ensuring a more organized flow and without risk of collapse. Fundamentally, this treatment proved to be a real and concrete tool to reduce morbidity and mortality. Randomized studies are needed to confirm the protocol’s potential to modify the natural course of the disease and support better resource management.

The great demand for hospital beds, which arises as a result of the rapid rise in the number of cases and long time to discharge already hospitalized patients, is capable of taking a health system to a critical level and failure. With the current epidemiology of the increase in hospitalizations for COVID-19, this alert serves as a strong impulse to study and evaluate the effectiveness of the pre-hospital approach with early reception, as soon as the symptoms start or at the first sign of persistence or worsening (8). The most certain approach for the disease is to treat it as early as possible. In some regression analysis, the timeto-treat was shown to be the most important predictor. It is important to consider that most patients with COVID-19 who arrive at the hospital through emergency medical services do not initially require advanced medical care (9). Once hospitalized, approximately 25% of patients progress to mechanical ventilation, advanced circulatory support or renal replacement therapy. Therefore, it is conceivable that some of the hospitalizations, if not most, could be avoided if patients were seen early on an outpatient basis, even with some severity criterion. Additionally, McCullough and collaborators (1) emphasize that COVID-19, as an infectious disease, is amenable to therapy at the beginning of its course and whose chances of response are less when this therapy is implemented late in hospitalized patients and in terminal stages.

The patients in this study had severity criteria, being classified as moderate (67.62%) or severe (32.38%). Among moderates, 71,12% had risk factors for severity, according to the WHO Clinical Management Guidelines (2). In the study by Richardson and collaborators (10), the risk factors most associated with mortality were hypertension, obesity and diabetes. In the present study, the presence of comorbidities can be seen in Figure 1, the most prevalent of which are in accordance with the literature (obesity - 46.67%, hypertension - 40.48% and diabetes - 14.76%).

We observed that the mortality of the study was lower than that of other case series of patients with criteria for hospitalization, with only 2 deaths (0,9%, Table 1). In a retrospective cohort study in China, Zhu F et al report that of the 191 patients included, 26% required care in the intensive care unit (ICU) and there was a mortality rate of 28% (11). A similar study conducted in the USA by Richardson S et al (10), showed that 14.2% of patients were treated in an intensive care unit, 12.2% received invasive mechanical ventilation and 21% died. Therefore, treatment with the protocol clearly demonstrated its potential to modify the outcome.

**Table 1.** Number of patients treated with the protocol *, recovered and deaths.

| Number of patients | N (%) |
| --- | --- |
| Total | 210 (100%) |
| Recovered | 208 (99,1%) |
| Deaths | 2 (0,9%) |

\*Prednisolone, enoxaparin, clarithromycin, axetylcefuroxime

Computed tomography of the chest (CT), although is not indicated as an isolated test of choice for diagnosis. Nevertheless, it is a valuable tool in the diagnostic aid, since it is useful in assessing the extent of the disease, in monitoring the evolution and detecting possible complications (12,33,44). During collapse, patients were diagnosed with COVID-19 by clinical and epidemiological criteria and the CT scan confirmed the pulmonary involvement of patients with progressive clinical worsening. The studies that evaluated the sequence of involvement on CT by COVID-19, indicate that in severe symptomatic patients, there is an evolution that seems common for cases that remained hospitalized. In the most initial phase, predominant ground-glass opacities occur, which are generally rounded and may be unilateral or bilateral. In this first phase, 20 to 30% of patients may already have consolidations. Some patients may already initially manifest a more serious disease, with ground-glass and / or diffuse consolidations (“white lung”), also indicating a poor prognosis. There were 27,3% of patients in this stage of extensive tomographic impairment in our sample.

This study raises the reflection that early reception, already with severity criteria, is fundamental and decisive in preventing fatalities, in addition to bringing immeasurable economic and psychosocial benefits, avoiding widespread panic.

Treating the inflammatory phase early, after the seventh day of symptoms and / or the presence of lung injury, has the potential not only to reduce mortality, but also to reduce the risk of morbidity, intubation and iatrogenesis (15,16,17,18). This measures are supported by the pathophysiology and also by the ethical principle of secondary prevention. This is defined as early action to intercept or halt the evolution of an already established disease. This applies to the early identification of the risk of an unfavorable outcome and implementing timely treatment to reduce sequelae, limitations and disabilities (19).

Publications showing the benefit of corticosteroid therapy in reducing hospital stay, intubation and deaths are crucial to corroborate the potential of this therapy in critically ill patients. Additionally, the present study highlights that good results observation of corticosteroid therapy before hypoxemia, at the beginning of phase II, may expand its indication and reinforce the hypothesis that the disease course may be changed earlier.

The fear of increasing viral replication with early use of corticosteroid therapy has not been confirmed, once an inflammatory phase has been established, a time when viral replication is no longer prevalent, and everything indicates that the development of a competent immune response has already been established. Documenting the SARS-COV-2 specific cellular immunity of patients with initial lung injury is essential, as it would demonstrate the absence of clinical worsening due to the low risk of increased viral replication explaining the success of treatment appropriately.

We believe that enoxaparin was decisive in controlling thrombogenesis considering the pathophysiology of the disease, as well as the modulation of immunity with the use of macrolides. However, studies are needed to confirm the magnitude of action of this drug in the management of COVID-19. Several studies are underway to determine the role of anticoagulants in the treatment of COVID19 due to the findings at autopsies.

Our experience with the collapse of Pará overflowed the solidarity spirit of the Pará’s doctors, as well as the infinite capacity of the human mind to solve problems in the face of adversity. There were more than 500 patients treated with this protocol in the period of the health system collapse and only 2 registered deaths. From this series, data from 210 patients were auditable. The experience of treating patients in the pulmonary phase of COVID-19 on an outpatient basis was widely disseminated among doctors in Belém and, since then, the treatment of these patients has remained primarily on an outpatient basis, even with hospital beds available after the pandemic has slowed down its pace. Thus, this therapeutic approach makes hospitalization an exception and the evolution to death is increasingly unlikely.

The authors suggest that patients with COVID19 symptoms be submitted to Chest CT Scan, d-dimer, lactate dehydrogenase and complete blood count, when seen for the first time. The same should be done on symptoms day 7, and if positive results, with ground-glass pattern, are found on CT Scan, the protocol should be introduced on an outpatient basis.

## CONCLUSION

1. The use of this therapeutic regimen as the first choice for COVID-19 pneumonitis will dramatically decrease the evolution of pneumonia, reducing the need for hospitalization and the use of respirators, favoring the health system and preventing fatalities.
2. This experience stimulates the study of the possible correlation between the development of competent cellular immunity in synchrony with the development of pulmonary inflammation, since there were no signs of viral replication or worsening of the disease.
3. It also opens the way to create a study with outpatient monitoring for COVID-19 initial pneumonitis, preventing unnecessary hospitalization. We still need to create this protocol with careful criteria for daily surveillance of patients. Suggesting that COVID-19 can become an ambulatory disease for most patients.
4. Emphasizes the importance of early reception for the treatment of initial pulmonary inflammation.
5. The data shown in this paper supported by the low mortality enforces that patients with initial COVID19 symptoms must be submitted to complete blood count, lactate dehydrogenase and d dimer levels, in addition of CT SCAN of the chest on their initial visit and on the seventh day of symptoms. Ground-glass findings on the latest CT SCAN support the suggested protocol adoption, with high probability of success.

## Data Availability

NA

